# Disability Inclusion in National Surveys

**DOI:** 10.1101/2024.07.02.24309837

**Authors:** Caroline Cerilli, Varshini Varadaraj, Jennifer Choi, Fiona Sweeney, Franz Castro, Scott D. Landes, Bonnielin K. Swenor

## Abstract

National surveys are important for understanding the disparities that disabled people experience across social determinants of health; however, limited research has examined the methods used to include disabled people in these surveys. This study reviewed nationally representative surveys administered by the Centers for Disease Control and Prevention (CDC) and the U.S. Census Bureau that collected data in the past five years and sampled adults ≥18 years. Data from both publicly available online survey documents and a questionnaire emailed to survey administrators were used to determine whether surveys: 1) oversampled disabled people; 2) had a data accessibility protocol to support data collection; and 3) provided multiple data collection modalities (e.g., phone, paper). Of the 201 surveys identified, 30 met the inclusion criteria for the study. Of these 30 surveys, one oversampled disabled people, none had a data collection accessibility protocol, and 21 provided multiple data collection modalities. This study highlights barriers and opportunities to including disabled people in national surveys, which is essential for ensuring survey data are generalizable to the U.S. population.

## Introduction

Nationally representative surveys are essential for public health surveillance in the United States (U.S.). Survey data are especially important for identifying inequities impacting marginalized populations such as people with disabilities, who make up nearly 27% of U.S. adults living in the community.(1–3) Data from national surveys provide information about the current state of health in the country, determine priorities to address disparities, and guide the allocation of resources.(4) Survey data have demonstrated that disabled people face disparities across social determinants and drivers of health including healthcare, (5, 6) education,(7) housing,(8) and criminal justice (9, 10). Furthermore, disabled people who are multiply marginalized are more likely to experience health inequities.(5, 11, 12) The recent designation of disabled people as a health disparities population by the National Institutes of Health(13) elevates the need for survey data to include and be representative of the disability community in order to identify and address the health inequities impacting this population.

Despite the importance of data, existing literature suggests that disabled people are often excluded from research studies. Across fields of research, inaccessible study materials create barriers to recruitment, informed consent, participation, and retention of disabled people in research studies.(14) Disability is often used as an exclusion criterion for study participation in clinical trials and biomedical research, but often these exclusions are not scientifically justified.(15) Notably, one study found that out of 97 clinical trials, 68% excluded people with psychiatric disabilities, 42% excluded people with cognitive or intellectual disabilities, and 33% excluded people with visual impairment.(16)

Despite this data on the exclusion from research studies, the exclusion of disabled people from national surveys is understudied. Existing national survey questions used to measure disability (17–19) and current survey methodology have limitations that impact generalizability for disabled people.(19–21) Although more than 1 in 4 American adults has a disability,(3) this population has been described as a rare or hard-to-reach population within surveys. (22–25) Therefore, survey sampling methodology must seek to oversample disabled people. (24) Failure to oversample disabled people results in insufficient sample sizes for analysis of outcomes for disabled people overall, and per specific disability status (e.g., vision, hearing, mobility). It is also necessary that the strategies used to field surveys are accessible, meaning they are offered in a manner that permits disabled people the ability to fully participate in and complete the survey. (26) Failure to use universal design approaches that maximize the accessibility of surveys biases the sample of disabled people to primarily include people whose disability does not limit their ability to participate. (27, 28)

To improve understanding of the representativeness of national survey data, this study examines the sampling and data collection methodologies of nationally representative surveys administered by the Centers for Disease Control and Prevention (CDC) and the U.S. Census Bureau that assess health or social determinants of health.

## Methods

The data in this study did not involve human subjects research, and therefore did not require Institutional Review Board approval.

### Survey Selection

From May to June 2023, two researchers (CC and VV) independently reviewed 201 surveys administered by the CDC and the U.S. Census Bureau for inclusion in this study. The CDC and Census Bureau surveys were chosen as they both administer multiple nationally representative surveys in the U.S. and they include data and on health equity and social determinants of health across sectors.

We applied the following criteria for inclusion of surveys in this study: 1) described as nationally representative 2) fielded within the past five years (2018–2023), 3) sampled at the person and/or household level, and 4) sampled adults ≥18. Longitudinal surveys that primarily sampled respondents who were under age 18, even if followed into adulthood, were excluded. All surveys included in the study were administered in English; for surveys administered in English and Spanish, only the data from the English version were included.

### Survey Methodology Analysis

From June to December 2023, researchers JC and FS independently assessed the publicly available online information published by the survey administrator (CDC or Census Bureau or both) of each survey included in the study. Data was doubly abstracted for any mention of disability from survey websites and the publicly available survey documents linked to that website, such as reports, survey methodology, or sampling frame information. A third team member (CC) adjudicated any discrepancies in the data in collaboration with JC and FS.

Online survey methodology documents and information were used to collect data on survey oversampling of disabled people. In instances where conflicting information was found, researchers relied on the most recent documentation and information, indicated by date. Data were also collected on the survey data collection modalities used, including online forms, paper, in-person, and telephone. Surveys that utilized Computer-Assisted Telephonic Interview (CATI) were categorized as telephone distribution and surveys that utilized Computer-Assisted Personal Interviews (CAPI) were categorized as in-person distribution.

Researchers (JC, FS, CC) abstracted and coded information regarding accommodations for disabled people during survey data collection processes (e.g., large text questionnaires, American Sign Language (ASL) interpreter availability, or accessible facilities). These data were used to categorize surveys as having: 1) a data collection accessibility protocol, 2) any accommodations available for data collection, but not a data collection accessibility protocol, or 3) no mention of accommodations. Having a data collection accessibility protocol was defined by the presence of a plan or process designed to include disabled people in survey data collection that aligns with federal legal requirements, including the ADA and sections 504 and 508 of the Rehabilitation Act.(29, 30) If a survey indicated that an office or department was available to assist with accommodations but did not meet the criteria of having a data collection accessibility protocol, it was coded as having available accommodations only. Notably, surveys that indicated offering multiple distribution modalities but no other accommodations were considered to have no mention of accommodations.

### Survey Administrator Contact

From October to November 2023, researchers JC and FS sent a questionnaire to survey administrators inquiring if disabled people were oversampled and if accommodations were made available in the surveying process to supplement the publicly available information. Standardized questions were sent via email when a contact address was available on the survey website, or by online form if an email was not available. Once a week for three weeks after the initial contact, researchers sent a reminder message to survey administrators who had not yet responded.

The questionnaire asked: (1) “does the sampling frame or sampling population for this survey/study specifically include people with disabilities? If so, please provide links to that information and any related protocol documents”; (2) “does this survey oversample people with disabilities? For example, people who have difficulty with vision, hearing, mobility, and cognition. If so, please provide links to that information and any related protocol documents”; (3) “does your survey administration and data collection strategy include the provision of any accommodations for people with disabilities? For example, large text questionnaires, ASL interpreter availability, accessible facilities, or plain language documents. If so, please provide links to that information and any related protocol documents.” Responses from each survey were coded as “yes” or “no” for each question.

All data were categorical and analyzed using frequencies and percentages. The data was visualized using bar charts. Analyses were conducted using Microsoft excel Version 16.86.

## Results

Researchers identified 201 CDC and U.S. Census Bureau surveys. A total of 30 surveys met the selection criteria for the study; eight from the CDC, 14 from the U.S. Census Bureau, and eight jointly administered by both (**Exhibit 1**). A total of 171 surveys were excluded as they did not meet one or more of the inclusion criteria described above. Survey administrators from 20 of the 30 surveys included in the study responded to the questionnaire.

**EXHIBIT 1 table.** Surveys included for analysis. Source/Notes: SOURCE [Authors’ analysis.] NOTES [Inclusion criteria: (1) national in scope (as opposed to hosted exclusively in one state or region), (2) fielded within the past five years (2018–2023), (3) sampled at the person and/or household level (as opposed to the institutional level or above), and (4) focused primarily on adults 18+. Accessibility protocol is defined as a data collection procedure that follows all federal standards for disability access and inclusion. Surveys that used at least two of the following in survey distribution were coded as having multiple data collection modalities: online form, paper form, in-person contact, or phone call.].]

| **Federal Survey Administrator** | **Title** | **Oversampled for Disabled People** | **Accessibility Protocol** | **>1 Data Collection Modalities** |
| --- | --- | --- | --- | --- |
| CDC | Behavioral Risk Factor Surveillance System | No | No | No |
|  | Health Information National Trends Survey | No | No | Yes |
|  | National Health and Nutrition Examination Survey | No | No | Yes |
|  | National HIV Behavioral Surveillance System | No | No | No |
|  | National Survey of Family Growth | No | No | No |
|  | Pregnancy Risk Assessment Monitoring System | No | No | No |
|  | The Medicare Current Beneficiary Survey | Yes | No | Yes |
|  | The National Survey on Drug Use and Health | No | No | Yes |
| Census Bureau | Ask U.S. Panel Pilot | No | No | Yes |
|  | Census Military Panel | No | No | Yes |
|  | Decennial Census of Population and Housing | No | No | Yes |
|  | Household Pulse Survey | No | No | Yes |
|  | Identity Theft Supplement | No | No | Yes |
|  | Medical Expenditure Panel Survey | No | No | No |
|  | National Sample Survey of Registered Nurses | No | No | Yes |
|  | National Survey of College Graduates | No | No | Yes |
|  | National Teacher and Principal Survey | No | No | Yes |
|  | National Training, Education, and Workforce Survey | No | No | Yes |
|  | Police-Public Contact Survey | No | No | Yes |
|  | Principal Follow-Up Survey | No | No | Yes |
|  | Teacher Follow-Up Survey | No | No | Yes |
|  | Telephone Point of Purchase Survey | No | No | No |
| CDC and Census Bureau | American Community Survey | No | No | Yes |
|  | American Housing Survey | No | No | Yes |
|  | American Time Use Survey | No | No | No |
|  | Consumer Expenditure Survey | No | No | Yes |
|  | Current Population Survey | No | No | Yes |
|  | National Crime Victimization Survey | No | No | Yes |
|  | National Health Interview Survey | No | No | No |
|  | Survey of Income and Program Participation | No | No | No |

### Survey distribution

A variety of survey modalities were identified based on publicly available online information and administrator questionnaires. Thirteen surveys collected data via online forms, 10 by paper, 16 in person, and 18 over the telephone. Twenty-one surveys indicated multiple formats were available (**Exhibit 2**).

**EXHIBIT 2 figure.**
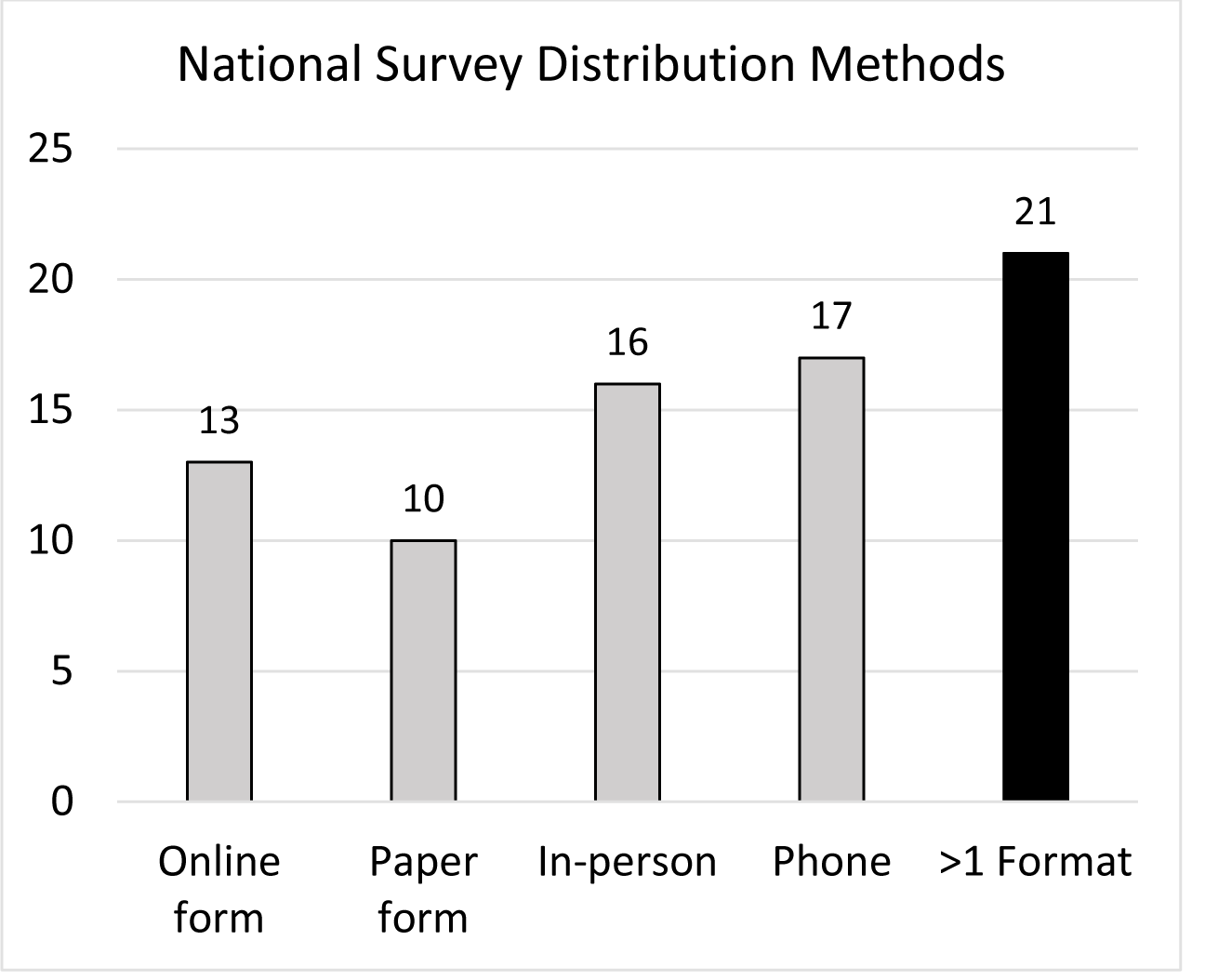
National survey distribution methods. Source/Notes: SOURCE [Authors’ analysis.] NOTES [Sample includes 30 national surveys from the U.S. Census Bureau and the Centers for Disease Control and Prevention that were described as nationally representative, fielded within 2018-2023, sampled at the person and/or household level, and sampled adults ≥18. The multiple formats bar indicates the number of surveys that used at least two of the following in survey distribution: online form, paper form, in-person contact, or phone call.].

### Oversampling

The Medicare Current Beneficiary Survey (MCBS) was the only survey with publicly available online methodology that indicated oversampling based on disability. However, of the survey administrators who responded to our questionnaire, none stated their survey oversampled based on disability, including the administrator from the MCBS (**Exhibit 3**).

**EXHIBIT 3 figure.**
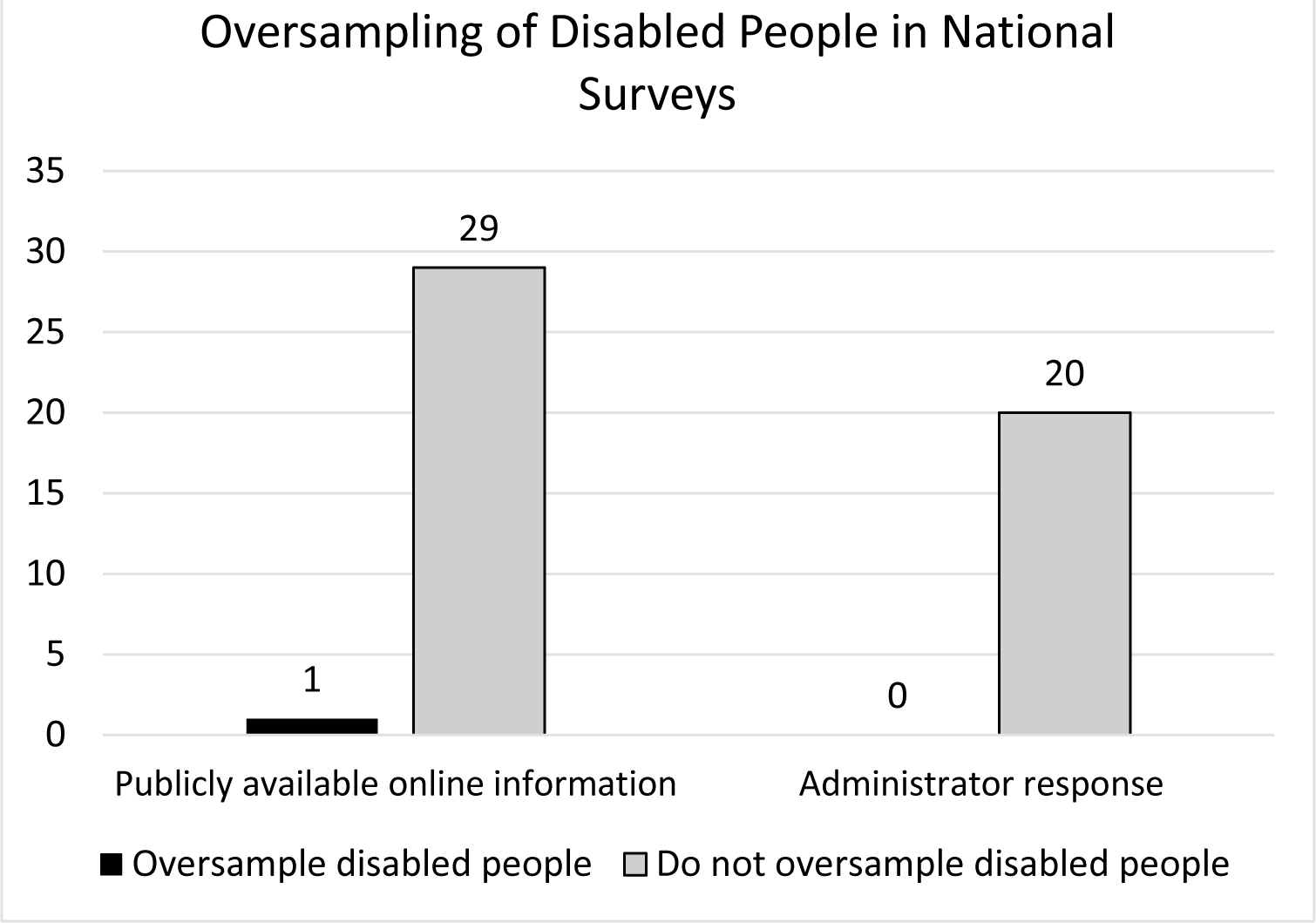
Oversampling of disabled people in national surveys. Source/Notes: SOURCE [Authors’ analysis.] NOTES [Sample includes 30 national surveys from the U.S. Census Bureau and the Centers for Disease Control and Prevention that were described as nationally representative, fielded within 2018-2023, sampled at the person and/or household level, and sampled adults ≥18. Twenty surveys’ administrator provided responses. Zero survey administrators reported oversampling disabled people.].

### Accommodations

None of the surveys publicly provided a protocol for providing accommodations to support data collection and survey participation. However, nine surveys indicated providing some form of accommodations for people with disabilities but no plan or process in place to ensure the accommodations include disabled people in survey data collection or to ensure data collection aligns with federal legal requirements. This includes two surveys that described accommodations in publicly available information and eight surveys that described accommodations in their questionnaire responses (**Exhibit 4**).

**EXHIBIT 4 figure.**
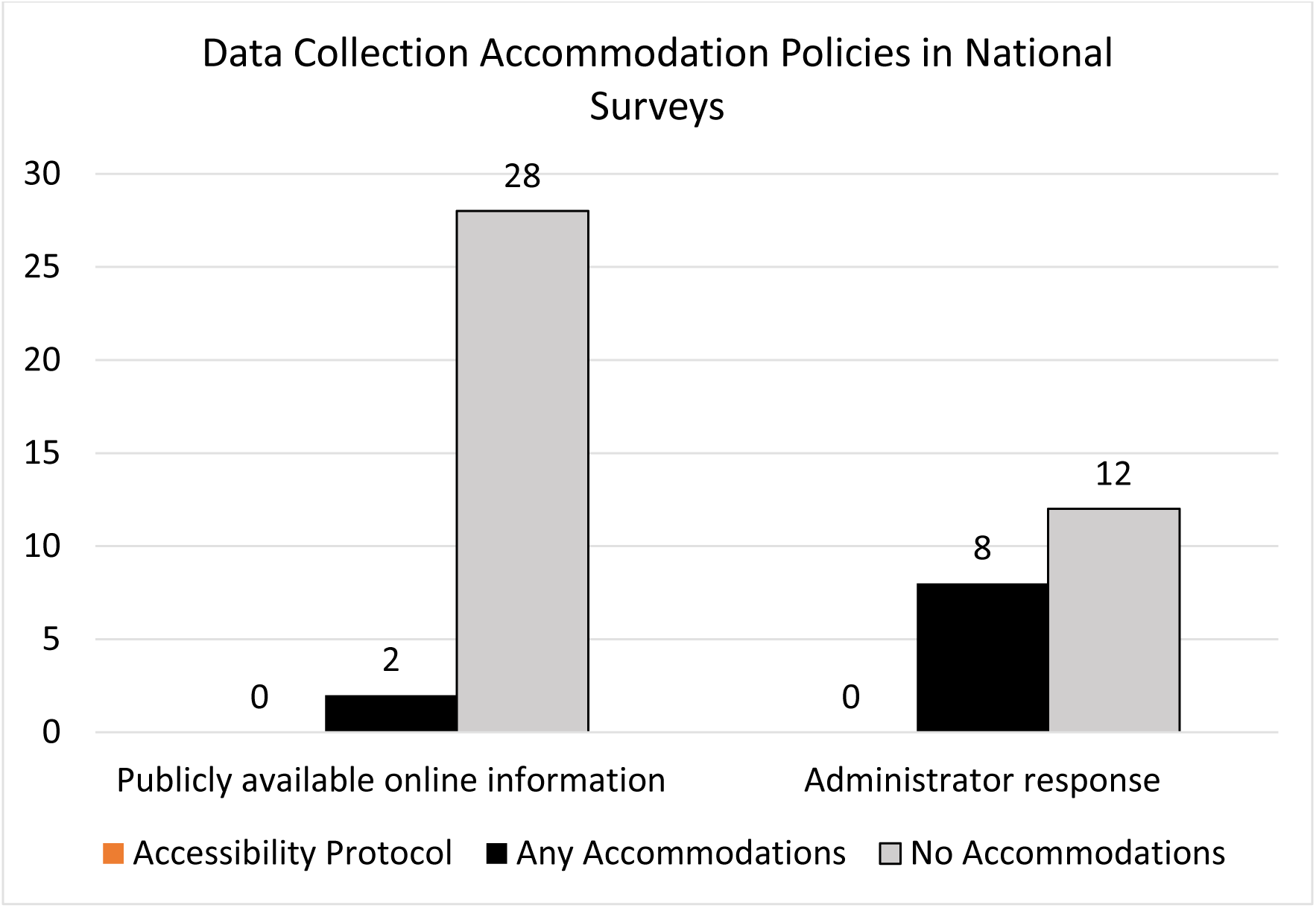
Data collection accommodation policies in national surveys. Source/Notes: SOURCE [Authors’ analysis.] NOTES [Sample includes 30 national surveys from the U.S. Census Bureau and the Centers for Disease Control and Prevention that were described as nationally representative, fielded within 2018-2023, sampled at the person and/or household level, and sampled adults ≥18. Twenty surveys’ administrator provided responses. Zero surveys had any publicly available information online or any administrator response that discussed an accessibility protocol, defined as a data collection procedure that follows all federal standards for disability access and inclusion.].

The National Health and Nutrition Examination Survey (NHANES) was one of two surveys that described accommodations in its publicly available information, but not a protocol to ensure accommodations support the inclusion of disabled people in data collection methods and meet federal legal requirements. The NHANES accessibility was limited to the in-person locations and presence of “a handicapped restroom with handrails, a hydraulic wheelchair lift, and a baby changing table.” Additionally, the National Survey on Drug Use and Health, a survey that collects data in person using a form, had public interviewer instructions which said to “use sensitivity and good judgement to determine whether [the respondent is] able to participate,” and to arrange the computer set up for data collection based on disability type. If needed, the interviewer is further instructed to manually place the respondents’ fingers on the appropriate computer keys if the respondent is unable to do so on their own. Deaf respondents are required to be able to read to participate.

Eight of the 20 administrator responses indicated accommodations are provided to participants, though typically on a case-by-case basis without a clear protocol. The National Sample Survey of Registered Nurses response stated, “Individuals requiring accommodation can reach out directly to survey administrator. Working with the project team, the request would then be addressed more specifically, possibly using the 508 Compliance office. … From there, the individual request is processed through the office based on need.”

Three administrators stated that participant’s use of a proxy may be allowed. The American Time Use Survey response stated that proxies could only be used “under special circumstances,” and the response from MCBS indicated proxies helped in 13% of their 2021 community-based interviews.

Five survey administrators responded that their surveys may provide ASL interpreters, though the majority could not guarantee that this service was available to all participants who needed it and was often limited by local availability. An administrator from the Consumer Expenditure Survey wrote, “Any ASL interpreters are up to Census availability, we unfortunately do not have any set procedure to make them available for respondents.”

Furthermore, some survey administrators were clear that accommodations were not provided, citing privacy concerns. An administrator from the National Survey of Family Growth wrote, “Our data collection does not make any provisions for disabled people. Given the sensitive nature of the NSFG content and the ancillary tools for the interviewer-administered mode of the survey (such as show cards and Life History Calendar), interviews must be conducted in private with no interpreters or proxies allowed, nor any people older than 4 years old present besides the respondent themselves. So face-to-face respondents must be able to see/read and hear well enough to be able to do the interview in private, and web respondents must be able to see/read well enough to do the survey on their device of choice.” Relatedly, an administrator from the Pregnancy Risk Assessment Monitoring Program wrote, “We do not provide accommodations for survey participants with disabilities. Also, PRAMS jurisdictions are reluctant to use Telecommunications Relay Services (TRS) due to PII concerns during data collection (e.g., respondents’ birth certificate information, sensitive health conditions, or risky health behaviors) since most would need to use a third-party contractor to perform these services.” Also, an administrator from the Principal Follow-Up Survey wrote, “The mailed questionnaires are not adapted for visual disabilities.”

## Discussion

These findings indicate that the design and data collection process used in U.S. federally administered national surveys may lead to the exclusion of disabled people. It is essential to ensure that disabled people are included and represented in federal national surveys data, because such survey data are used to inform policy and allocate resources. Data from the U.S. Census Bureau alone informs allocation of $2.8 trillion dollars, and the underrepresentation of disabled people in national surveys may result in insufficient funding to meet the needs of this population.(31) Failure to use survey strategies that ensures the full inclusion of disabled people in nationally representative surveys perpetuates structural ableism, or the systemic valuing of nondisability over disability.(32)

Recent policy has focused on improving disability representation in federal data and recognizes this is essential for advancing equity. President Biden’s Executive Order on Advancing Racial Equity and Support for Underserved Communities Through the Federal Government specifically calls for improved data collection on groups such as disabled people.(33) In response to this Executive Order, the Equitable Data Working Group was established to “outline a strategy for increasing data available for measuring equity and representing the diversity of the American people and their experiences.”(34) In March 2023, this working group published a progress report that stated, “To fully realize the potential of equitable data to drive better outcomes for all Americans, it will be especially necessary to create incentives and pathways for increased diversity and representation among public data practitioners and research community participants.”(35) Yet, these goals will remain unfulfilled until efforts are made to ensure disabled people can equitably participate in U.S. surveys.

A lack of accessible options for disabled people to participate in surveys was observed, suggesting that our national surveys are not meeting federal legal standards. Federal civil rights and regulations, such as the Rehabilitation Act of 1973, prohibit the exclusion of disabled people from federally funded programs and activities, including national surveys administered by the federal government, on the basis of disability.(30) However, none of surveys in this study described a data collection accessibility protocol for participants with disabilities.

Survey methods and protocols must develop protocols to maximize accessibility and adopt universal design approaches. One of the core principles of universal design is flexibility, which encourages multiple, equitable options to engage with information.(28) While this study found that 21 surveys include multiple distribution methods for data collection, further work is needed to examine if each of the modalities offered are accessible to and useable by disabled people. But even with the use of universal design, accommodations will be needed and are federally required. For example, Americans with Disabilities Act (ADA) specifically requires that national surveys such as those in this study provide all people with “effective communication,” which may include, but is not limited to, impartial ASL interpreters, large print, tactile communication, real-time captioning, or additional time to respond.(29, 36) However, our study suggests that U.S. national surveys are not compliant with the ADA. An administrator of the National Survey of Family Growth stated participants must see and hear “well enough” to complete their survey without accommodations, effectively excluding many people with sensory disabilities. Further, two survey administrator questionnaire responses indicated that they do not use interpreters for ASL users due to concerns of breaching health information confidentiality. Yet, the first tenant of the National Association of the Deaf Registry of Interpreters, a major organization that provides national ASL interpreter certification for the U.S., requires interpreters to adhere to confidentiality standards across all areas.(37) Additionally, many D/deaf ASL users strongly prefer using professional interpreters in clinical settings to any other mode of communication, and have stated that other methods, such as use of family proxy interpreters, impact confidentiality.(38, 39) This study also found just three surveys that mentioned use of proxy responses. Proxy responses are less reliable than self-report, and the direct perspective of a person with a disability should be prioritized. Therefore, accommodations should be made to collect the self-report wherever possible.(40)

Surveys purporting to be nationally representative must also consider oversampling disabled people. As more than a quarter of American adults are disabled,(3) it is striking that so few of the surveys included in our study oversampled for this population. The heterogeneity of the disability community should not be a deterrent to oversampling this population; heterogeneity is also present among racial and ethnic groups who are oversampled in the majority of U.S. surveys. Oversampling disabled people is necessary to support disaggregating disability data by specific disability status (e.g. vision hearing, physical disability), other demographic variables, and across geographic regions, which is essential for examining health inequities impacting multiply marginalized groups.

It is notable that we found inconsistencies in information collected from public survey information and survey administrator responses. For example, MCBS documents stated it does oversample on disability, but the survey administrator reported that it does not. While certainly survey protocols evolve and improve over time, transparent and consistent information is needed to support public trust and enhance use of the resulting data.

## Limitations

Data for this project are restricted to publicly available online information and questionnaire responses. It is possible that further information may exist that our team could not access. However, transparency in the design and methodology of federal surveys is critical to understanding if people from the disability community, as well as other underserved and underrepresented groups, are adequately represented. Further, this study was limited to surveys from the CDC and U.S. Census Bureau, and therefore not exhaustive of all federal surveys. Additionally, the surveys included primarily sampled noninstitutionalized adults. While collecting data from people living within institutions presents unique challenges, this exclusion has implications for the representation of disabled people who are more likely to be living in institutions, including long term care facilities and nursing homes.

## Conclusion

This study demonstrates the pervasive methodological barriers to the inclusion of disabled people in the design and data collection process of federally administered national surveys. The exclusion of disabled people violates federal policies, perpetuates structural ableism, and prevents opportunities for advancing equity, including health equity. Ensuring national survey data includes disabled people and is truly representative of the U.S. population will require collaboration across the federal statistical community, researchers, and policy makers and must become a national priority.

## Data Availability

All data produced in the present study are available upon reasonable request to the authors.

